# Sphingolipid and ceramide associations with tau pathology vary across diverse ethnoracial groups in postmortem brain tissue

**DOI:** 10.1101/2025.11.04.25339489

**Authors:** Annalise Schweickart, Richa Batra, Kevin Huynh, Collette Blach, Alexandra Kueider-Paisley, Joseph S. Reddy, Gregory Klein, David A. Bennett, Peter J. Meikle, Bin Zhang, Philip L. De Jager, Laura Heath, Jaclyn Beck, Jo Scanlan, Nicholas T. Seyfried, Dennis W. Dickson, Nilüfer Ertekin-Taner, AMP-AD Diverse Cohorts Working Group, Alzheimer’s Disease Metabolomics Consortium, Rima Kaddurah-Daouk, Matthias Arnold, Jan Krumsiek

## Abstract

Metabolic dysregulation is a hallmark of Alzheimer’s disease (AD), with numerous studies characterizing metabolic pathways associated with AD onset and progression. A significant limitation of these studies has been a predominant focus on non-Hispanic white participants. Despite evidence that AD prevalence, progression, and biomarkers differ across ethnoracial groups, it remains unclear whether previously identified metabolic dysregulation in AD brains generalizes across populations. We addressed this gap by analyzing large-scale metabolomics data from 547 postmortem dorsolateral prefrontal cortex brain tissue samples of Hispanic American, Non-Hispanic African American, and White subjects, providing the largest multiethnic AD brain cohort analyzed to date. A metabolome-wide association study examined how relationships between metabolite abundance and AD neuropathology varied by ethnoracial group. Sixty metabolites exhibited significant heterogeneity with tau pathology (Braak stage), with enrichment in tricarboxylic-acid-cycle intermediates, dipeptides, and sphingolipid-ceramide pathway lipids. These findings reveal ethnoracial-specific metabolic signatures of tau pathology and emphasize the need to evaluate emerging therapeutic targets across diverse groups.

## Introduction

Alzheimer’s disease (AD) is a major global health concern, affecting millions worldwide^1^. The prevalence of AD varies among different populations: recent estimates show 19% of non-Hispanic African Americans (NHAAs) age 65+ have Alzheimer’s dementia as compared to 14% in Hispanic Americans (HAs) and 10% in non-Hispanic Whites (NHWs)^2^ although these estimates vary^3,4^. These disparities in prevalence are assumed to arise from genetic, environmental, and socio-economic factors, including access to healthcare, education, and environmental stressors, as well as the underlying discrepancies in medical risk factors for AD, such as depression, diabetes, and hypertension^5,6^.

To obtain a better understanding of these ethnoracial disparities at the molecular level, recent AD research has focused on increasing representation of diverse populations in multi-omics studies^6–8^. Genome-wide association studies (GWAS) have identified that the strongest genetic risk for AD among NHWs, the *APOE4* allele, is more common in NHAAs^9^ and less common in certain HA populations^10,11^ compared to NHWs. Further, *APOE4* allelic dosage has been associated with faster decline on a test of verbal memory among NHWs but not among NHAAs, suggesting divergent biological mechanisms that contribute to disease progression^12^. Moreover, ethnoracial factors have been linked to distinct AD-related biomarkers, including lower levels of p-tau_181_ and Aß_40_ in the cerebrospinal fluid of African Americans^13^ and higher S100B in the serum of Mexican Americans^14^. Such differences in individual biomarker levels result in higher exclusion rates of diverse groups from clinical trials^15,16^, limiting the generalizability of trial results. Overall, these findings underscore the need to investigate the molecular mechanisms underlying AD across diverse populations to guide future research and pharmaceutical developments.

Variation in metabolic physiology may be an important driver of heterogeneity in AD manifestation and risk across ethnoracial groups. Metabolic diseases such as diabetes, obesity, and hypertension exhibit greater prevalence among NHAAs and HAs and confer different AD risk relative to NHWs^3^. Furthermore, metabolic factors, including fatty acid-binding protein (FABP), glucagon-like peptide-1 (GLP-1), and insulin, are differentially abundant in some HA populations compared to NHWs^15^. Metabolic dysregulation is a recognized characteristic of Alzheimer’s disease^17^, evidenced by altered brain glucose uptake detectable via fluorodeoxyglucose (FDG)-based positron emission tomography (PET) scans^18^, by the *APOE4* allele, which influences lipid metabolism^19^, and by metabolomics analyses showing metabolic changes in postmortem brain tissue correlated with AD pathology^20,21^. Metabolism has become a strategic focus in AD treatment, as reflected in the growing interest in therapies such as GLP-1 agonists for insulin and glucose regulation^22^ or dietary and lifestyle interventions to enhance cognitive health^23,24^. However, how metabolic differences across ethnoracial groups contribute to clinical and molecular heterogeneity in AD remains insufficiently characterized.

To our knowledge, this is the first study to apply untargeted metabolomics to a large multiethnic postmortem brain collection to date. In this study, to elucidate metabolic alterations associated with AD across different ethnoracial groups, we applied metabolomics to postmortem brain tissue samples from the Accelerating Medicines Partnership – Alzheimer’s Disease (AMP-AD) Diverse Cohorts Study (Figure 1). Metabolomics offers a powerful approach to reveal small-molecule changes reflecting both genetic and environmental influences on health. We applied high-performance liquid chromatography–tandem mass spectrometry (UPLC/MS-MS)-based metabolomics to profile ∼900 metabolites in the dorsolateral prefrontal cortex (DLPFC) of 547 donors, collected from four study sites across the United States. Donors were balanced across major ethnoracial groups commonly represented in the United States. Rigorous quality-control measures were implemented to remove site-related variation, followed by association testing using linear models with interaction analysis to identify metabolites differentially linked to AD pathology across ethnoracial groups. Our findings highlight several metabolic pathways that exhibit heterogeneity across these groups, underscoring the importance of considering population-specific metabolic influences in AD research.

**Figure 1:**
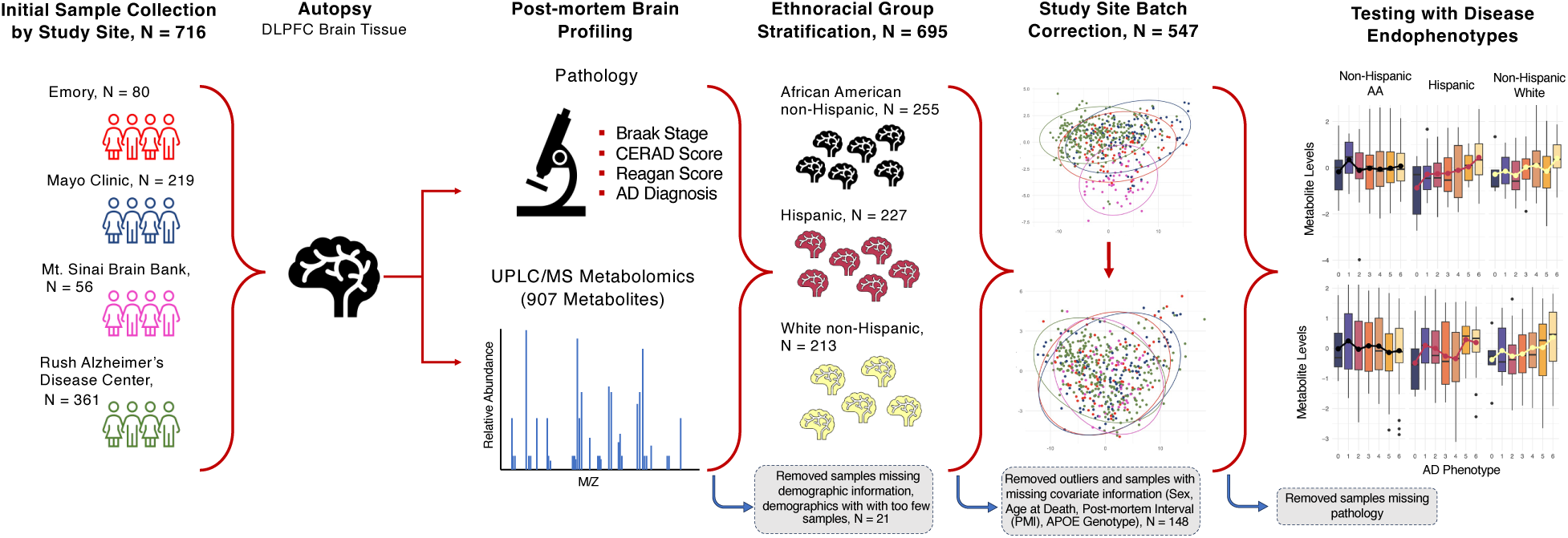
Study Overview.

## Methods

### Brain sample and clinical metadata collection

The metabolomics data used in this study originates from the Accelerating Medicines Partnership – Alzheimer’s Disease (AMP-AD) Diversity Cohorts Study^7,25^, a collaborative project involving multiple study sites and designed to capture a broad spectrum of multi-omics data, including proteomics, genomics, and metabolomics from brain tissue samples. 741 brain samples from the temporal cortex and dorsolateral prefrontal cortex were collected from 716 subjects across four study sites—Mayo Clinic, Rush Alzheimer’s Disease Center, Mount Sinai University Hospital, and Emory University—with a focus on enriching for samples from African American and Hispanic American populations.

Demographic and biological variables available across studies included sex, age at death, postmortem interval (PMI), and *APOE* genotype. Self-reported race (American Indian or Alaska Native, Asian, Black or African American, White, Other) and self-reported ethnicity (“is Latin American/Hispanic” yes or no) were also collected. Three ethnoracial groups had sufficient sample size for further analysis and were defined as follows: all subjects who responded “yes” to “is Latin American/Hispanic” were labeled as “Hispanic American” (HA). Of the remaining non-Hispanic population, those who reported their race as “White” were labeled as “non-Hispanic White” (NHW) and those who reported their race as “Black or African American” were labeled as “non-Hispanic African American” (NHAA).

AD likelihood, diagnosis, and neuropathological burden were assessed using semi-quantitative neuropathological measures without knowledge of clinical information: Braak staging was used to classify the severity of tau pathology in the brain, ranging from stages I–VI^26^. For non-Mayo Clinic samples, CERAD values were used to assess the severity of amyloid pathology on a four-level scale, categorized from frequent neuritic plaques (CERAD 1) to no pathology (CERAD 4)^27^. Moreover, NIA-Reagan criteria a combination of Braak staging and CERAD scores, were categorized from high likelihood (Reagan score 1) to no AD (Reagan score 4)^28^. For Mayo Clinic samples, Thal phase was used to measure amyloid. Finally, a binary AD diagnosis was derived using the following criteria: AD case status was assigned where Braak stage was ≥ 4 and CERAD score was ≤ 2; control status was assigned where Braak stage was ≤ 3 and CERAD score was ≥ 3. Samples meeting neither case nor control criteria were were excluded from downstream analyses requiring AD diagnosis. All Mayo Clinic AD samples had Braak neurofibrillary tangle (NFT) stage of ≥ IV and evidence of Thal≥ 2 amyloid deposits and were given diagnosis of AD according to the NINCDS-ADRDA criteria^29^.

For the present study, samples from brain regions other than dorsolateral prefrontal cortex (DLPFC) were removed (25 samples), as were samples with missing ethnoracial information or belonging to ethnoracial groups with less than 15 subjects (21 samples). Samples missing key covariate information, including sex, age at death, PMI, and *APOE* genotype were also removed (146 samples). Detailed descriptions of study sites, sample collection and phenotype harmonization have been previously published^30^.

### Metabolomics data acquisition

Brain tissue samples (n = 741) and NIST plasma samples (n = 44) were processed using Metabolon’s global metabolomics platform. Upon receipt of brain tissue, samples were accessioned into a laboratory information management system (LIMS), stored at −80°C, and prepared using an automated MicroLab STAR® system. Metabolite extraction involved protein precipitation with methanol, followed by centrifugation. Each extract was split into four aliquots and profiled by UPLC-MS/MS using two C18 reverse-phase positive-ion runs (early- and late-eluting), one C18 reverse-phase negative-ion run, and one HILIC negative-ion run. Peak identification was conducted using Metabolon’s proprietary software, referencing a library of authenticated standards and recurrent biochemicals. Data normalization corrected for inter-day instrument variability. Quality control included internal standards, pooled matrix samples, and process blanks to ensure data reliability. A total of 1,388 metabolites were identified and reported.

A detailed description of the metabolomics measurement procedures is provided in **Supplementary Material 1.**

### Data preprocessing and study site correction

Metabolites with more than 25% missing values were removed (481 metabolites). Probabilistic quotient normalization^31^ was applied to correct for sample-wise variation, followed by log_2_ transformation. Remaining missing values were imputed using a k-nearest-neighbor-based algorithm (kNN)^32^. The local factor outlier (LOF) method^33^ was used to identify sample outliers (2 samples). To account for irregularly high or low individual concentrations, values with absolute abundance above *q = abs(qnorm(0.0125/n*)) under a scaled-data assumption, with *n* representing the number of samples, were set to missing. This formula finds cutoffs to flag values with less than 2.5% two-tailed probability to originate from the same normal distribution as the rest of the measurement values, after applying a Bonferroni-inspired correction factor (division by sample size). These new missing values were then imputed by another round of the k-nearest-neighbor algorithm. After preprocessing, the final data matrix consisted of 907 metabolites measured for 547 samples. All preprocessing was performed using the maplet^34^ R package.

To adjust for site variability on metabolite abundance while preserving true biological signal, we employed ComBat, a parametric empirical Bayes method for batch effect correction, where batch in this case is the study site^35^. In brief, ComBat assumes that the observed abundance for metabolite *m* in sample *i*, denoted *Y*_*i*,*m*_, can be decomposed into a biological component plus a batch-specific effect:

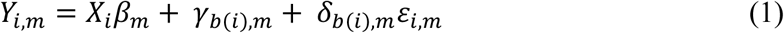

where *X*_*i*_ is a vector of our covariates of interest for sample *i* (sex, age at death, PMI, *APOE* genotype, and ethnoracial group), *β*_*m*_ is the corresponding coefficient vector for metabolite *m*, *b(i)* represents the batch (in our case, study site) membership of sample *i*, and *γ*_*b*(*i*),*m*_ and *δ*_*b*(*i*),*m*_ model the mean and variance adjustments for each batch, respectively. ComBat pools information across metabolites to shrink these batch-specific parameters (*γ*_*b*(*i*),*m*_and *δ*_*b*(*i*),_) toward global estimates via empirical Bayes.

*X*_*i*_was included in the batch-correction step due to observed imbalances in the distribution of ethnoracial groups and age at death across study sites. The metabolite variation due to covariates designated in *X*_*i*_ remains preserved so that true biological variability of interest is retained after correction.

To ensure this correction step correctly removed the study site signal while preserving the signal of the covariates in *X*_*i*_, a variance partitioning analysis was performed^36^. The following linear model was run for every metabolite:

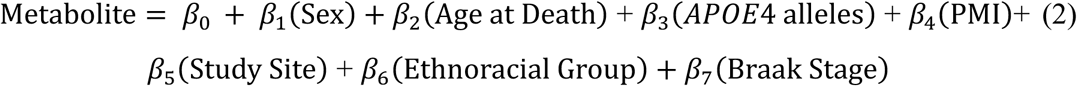

The variance explained for each of the model variables was collected for each metabolite and compared between pre- and post- correction.

### Statistical analysis

After correcting for study site variation, we performed association analysis between metabolites and the four AD-related neuropathological measure, Braak stage, CERAD score, Reagan score, and AD diagnosis. For each measure (outcome) and each metabolite, the following models were calculated:

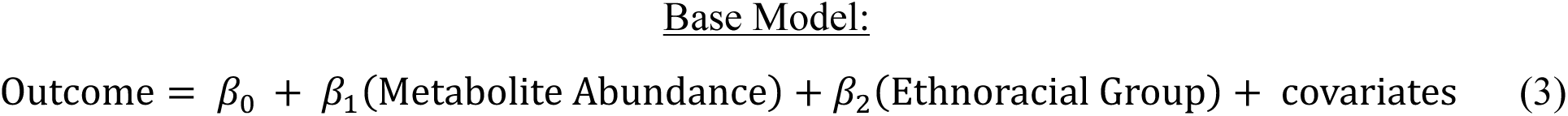

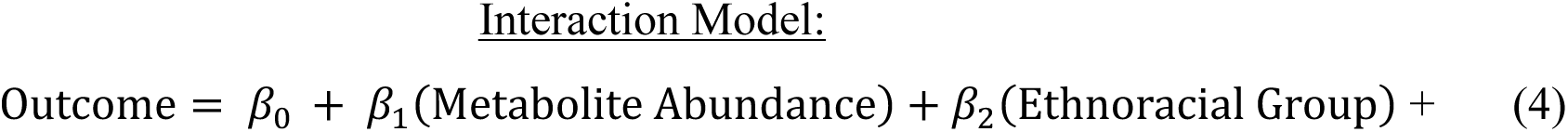

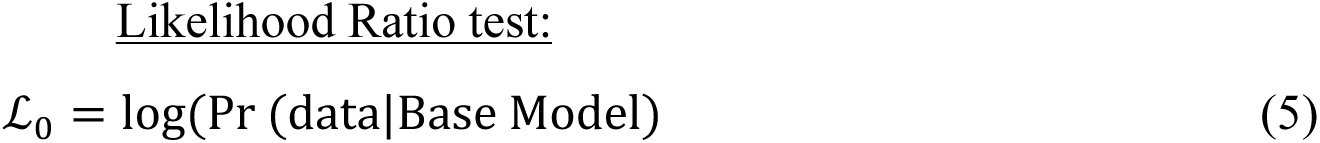

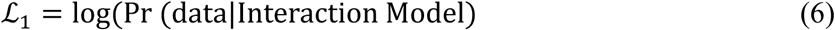

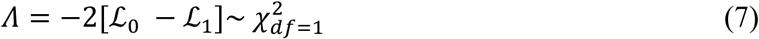

Since Braak, CERAD, and Reagan are ordinal parameters, an ordinal regression model was used for both base and interaction models using the rms package in R^37^. For AD diagnosis, a logistic regression model was used. Covariates included sex, age at death, PMI, and number of *APOE4* alleles. The likelihood ratio test between the two regression models with (4) and without (3) the interaction term of metabolite and ethnoracial group assesses the added value of the interaction term. Specifically, ℒ_0_ (5) represents the log-likelihood of the base model and ℒ_1_ (6) represents the log-likelihood of the interaction model. The likelihood ratio statistic 𝛬, calculated in (7), approximately follows a Chi-square distribution, which was used to calculate a p-value of the interaction. To account for multiple hypothesis testing, p-values were corrected using the Benjamini-Hochberg (BH) method^38^.

For metabolites with significant interaction effects, we additionally examined each ethnoracial group separately to determine which groups were contributing to the observed heterogeneity. In these within-group analyses, we used the same base model (3) without the ethnoracial group covariate. This allowed us to identify which ethnoracial group (if any) had a significant metabolite-outcome association and thereby pinpoint the source of the interaction signal.

## Results

### Sample characteristics

This study examined how associations between metabolites in the dorsolateral prefrontal cortex (DLPFC) and AD neuropathology differ across ethnoracial groups. Four neuropathological measures were analyzed: Braak stage, CERAD score, Reagan score, and AD diagnosis (derived from Braak and CERAD scores or Thal phase). The analysis was restricted to samples with complete data for all covariates (sex, age at death, PMI, *APOE4* allele count) and neuropathology. Detailed characteristics of the unfiltered data with 716 DLPFC samples can be found in **Supplementary Table 1**. Of the 547 preprocessed brain tissue samples, Braak stage information was available for the largest number of subjects, resulting in a study group of 545 subjects (**Table 1**), while CERAD and Reagan score analyses included 428 samples and AD diagnosis analysis included 426 samples. Samples used for the Braak analysis consisted of 220 non-Hispanic African Americans (NHAA), 148 Hispanic Americans (HA), and 177 non-Hispanic Whites (NHWs). Among these 545 participants, 59% had a neuropathologic diagnosis of AD, 65% were female, and 38% were *APOE4* allele carriers. Stratified by ethnoracial group, *APOE4* frequency was 52% in NHAAs, 30% in HAs, and 29% in NHWs. Detailed distributions of neuropathology measures across ethnoracial groups can be found in **Supplementary Table 2.**

**Table 1.** Descriptive characteristics of study participants, tabulated by ethnoracial group.

| Characteristic | Overall<br>N = 545<sup>1,2</sup> | Non-Hispanic<br>African American<br>N=220<sup>1</sup> | Hispanic<br>American<br>N = 148<sup>1</sup> | Non-Hispanic<br>White<br>N=177<sup>1</sup> |
| --- | --- | --- | --- | --- |
| Study Site |  |  |  |  |
| Emory | 77 (14%) | 40 (18%) | 0 (0%) | 37 (21%) |
| Mayo Clinic | 117 (21%) | 29 (13%) | 73 (49%) | 15 (8.5%) |
| Mt Sinai Brain Bank | 55 (10%) | 28 (13%) | 27 (18%) | 0 (0%) |
| RADC<sup>3</sup> | 296 (54%) | 123 (56%) | 48 (32%) | 125 (71%) |
| Sex |  |  |  |  |
| Female | 356 (65%) | 141 (64%) | 88 (59%) | 127 (72%) |
| Male | 189 (35%) | 79 (36%) | 60 (41%) | 50 (28%) |
| Number of APOE4 alleles |  |  |  |  |
| 0 | 336 (62%) | 106 (48%) | 104 (70%) | 126 (71%) |
| 1 | 168 (31%) | 91 (41%) | 35 (24%) | 42 (24%) |
| 2 | 41 (7.5%) | 23 (10%) | 9 (6.1%) | 9 (5.1%) |
| Alzheimer's Disease Diagnosis |  |  |  |  |
| Case | 321 (59%) | 126 (57%) | 90 (61%) | 105 (59%) |
| Control | 104 (19%) | 46 (21%) | 14 (9.5%) | 44 (25%) |
| Other<sup>4</sup> | 120 (22%) | 48 (22%) | 44 (30%) | 28 (16%) |
| Postmortem Interval (hours) | 8 (6, 14) | 8 (6, 15) | 8 (5, 15) | 7 (5, 10) |
| Age at death (years) | 84 (76, 90) | 82 (76, 89) | 81 (73, 88) | 90 (83, 95) |

<sup>1</sup>n (%); Median (Q1, Q3)
<sup>2</sup>Sample size after removing missing Braak pathology
<sup>3</sup>RADC: Rush Alzheimer's Disease Center
<sup>4</sup>Did not meet case or control criteria (Case: Braak stage $\geq 4$ and CERAD score $\leq 2$ ; Control: Braak stage $\leq 3$ and CERAD $\geq 3$ )

Notably, the NHW samples had a higher age at death as compared to HAs and NHAAs. In addition, ethnoracial groups were not equally represented in all study sites, as the Emory site contributed no Hispanic American samples, and the Mt. Sinai site contributed no NHW samples (**Table 1**).

### Metabolomic profiling and batch effects

Brain metabolomic profiles were measured using Metabolon Inc.’s untargeted metabolomics platform. This platform identified 1,388 metabolites from various chemical classes (super-pathways) in the DLPFC tissue samples, which were filtered down to 907 metabolites with sufficient coverage across samples **(Supplementary Data 1).**

Importantly, 70% of the metabolites were significantly associated with study site (FDR adjusted p-value < 0.05, **Figure 2a**). To address this unwanted variation introduced by study site, which could obscure the biological signals of interest, the batch correction method ComBat^35^ was applied. This method is able to remove batch effects despite the main biological variable (ethnoracial group) being confounded by the batch variable (site). For details, see Methods. Variance partition analysis^36^ (VPA) showed that ComBat successfully removed unwanted variation from the study site while preserving the desired variation from variables of interest (**Figure 2b**). The variation associated with study site decreased from an average of 4.2% per metabolite before correction to 0.2% after correction, while variation linked to all other variables of interest, including ethnoracial group, remained largely unaffected.

**Figure 2.**
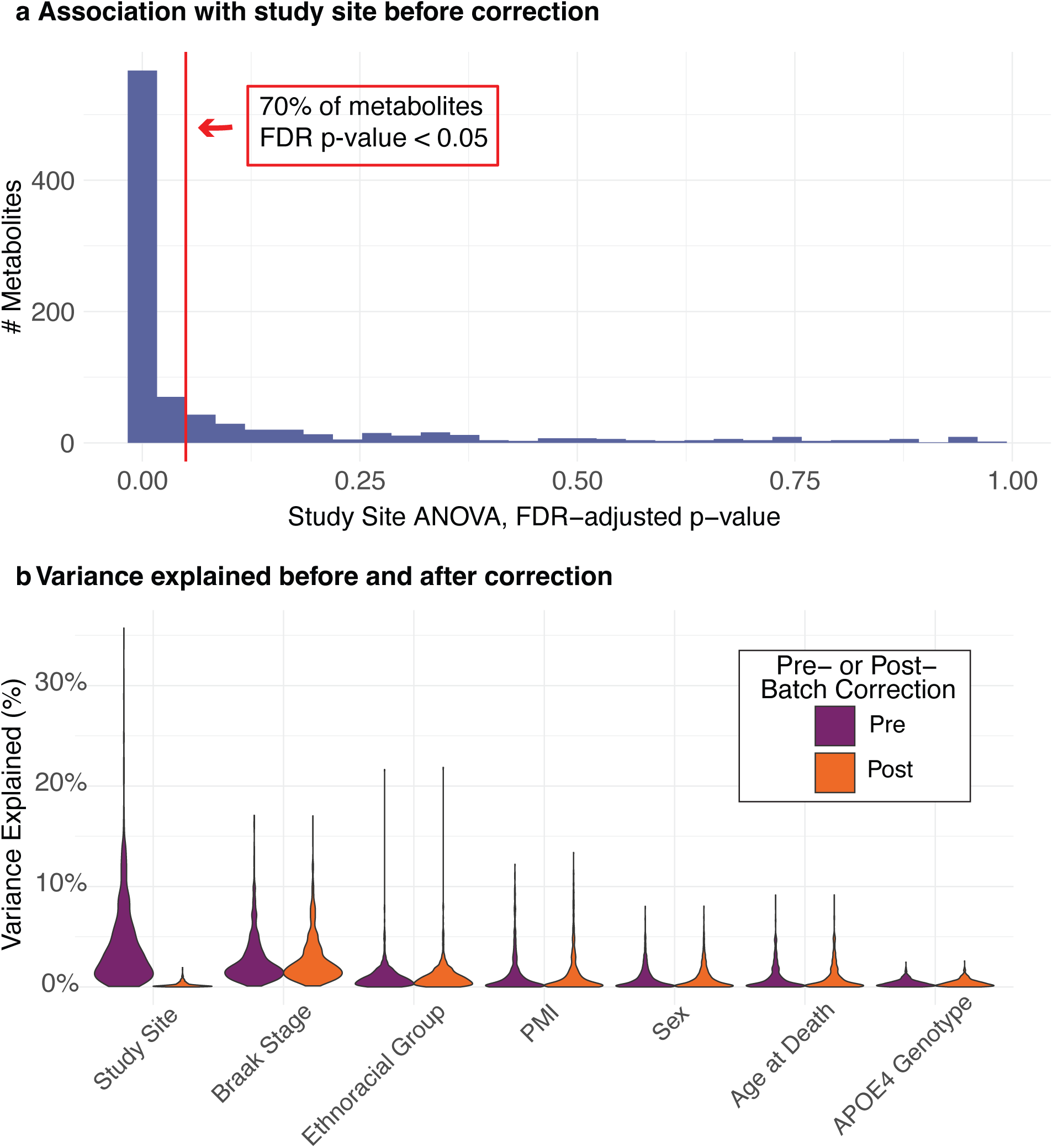
QC and batch correction. **a** 70% of the metabolites were significantly associated with the sites (FDR adjusted p-value < 0.05), indicating strong batch effects. **b** ComBat was used to remove batch effects in the presence of confounding between batch variable (study sites) and biological variables of interest. Variance partition analysis1showed that variation associated with study sites decreased from an average of 4.2% per metabolite before correction to 0.2% after correction, while variation in all other variables remained largely unaffected.

### Ethnoracial interaction screen identifies 60 Braak-stage-dependent metabolites

Out of 907 metabolites used in this analysis, lipids made up the largest category (45.5%), followed by amino acids (19.2%), nucleotides (6.5%), carbohydrates (5.0%), cofactors/vitamins and peptides (each 4.3%), xenobiotics (3.6%), energy-related metabolites (1.1%), and an additional ∼10% that remain uncharacterized (**Figure 3a, left**).

**Figure 3.**
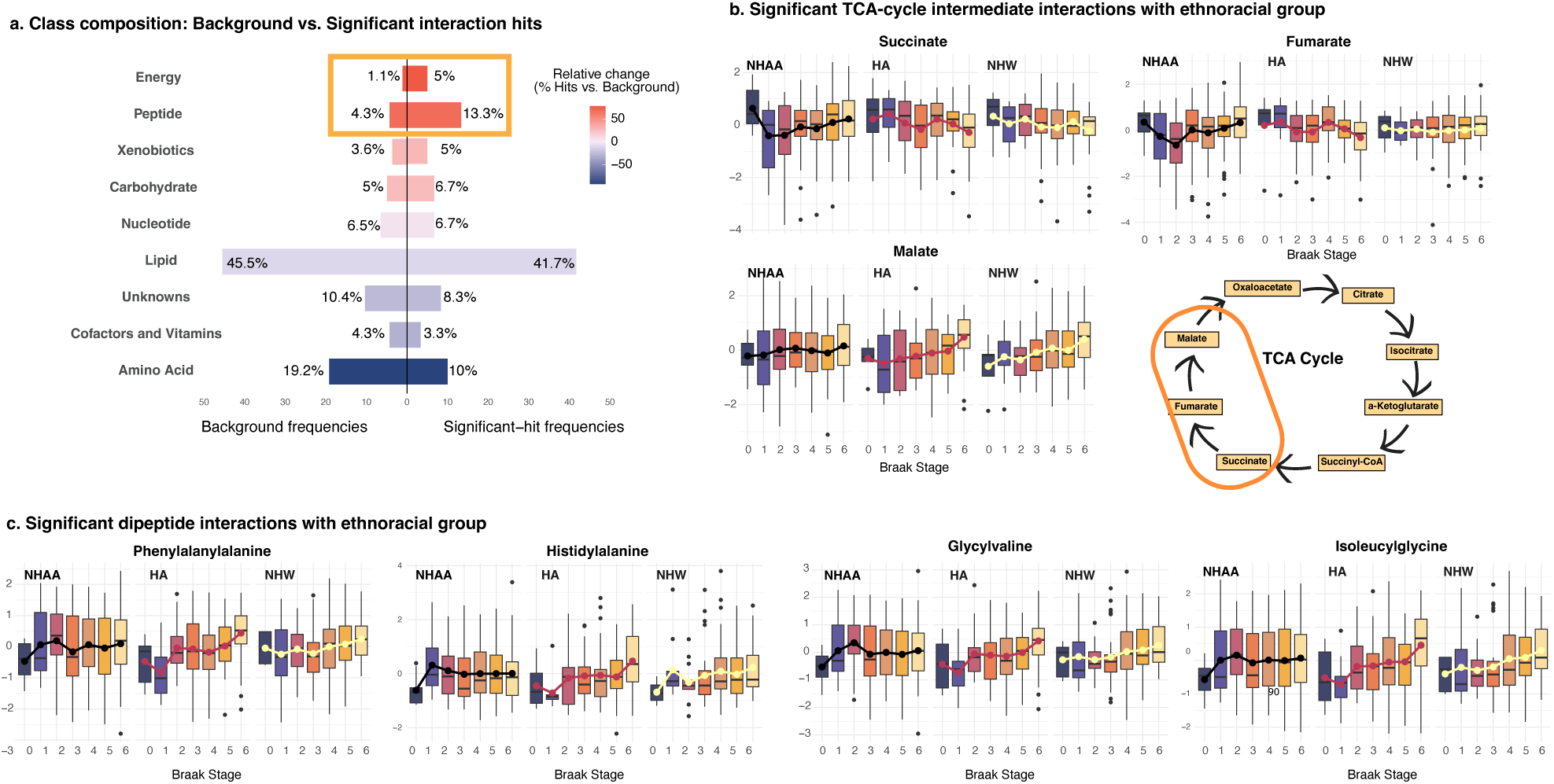
Ethnoracial differences in the relationship between brain metabolite abundance and Braak neurofibrillary-tangle stage. **a** Among 907 quantified metabolites, 60 showed an interaction between Braak stage and ethnoracial group at FDR < 0.10. The distribution of these heterogeneous hits (right) largely mirrors the overall chemical profile (left), though dipeptides and energy-pathway metabolites are significantly proportionally enriched. **b** Key tricarboxylic-acid-cycle intermediates (malate, fumarate, succinate) also diverge by ethnoracial group: abundances increase with Braak stage in Hispanic Americans (HA), remain flat or decline in non-Hispanic Whites (NHW), and show little change in non-Hispanic Africans American (NHAA). **c** Box plots for four significant dipeptides showed a marked, stage-dependent rise in HAs, a modest rise in NHWs, and no clear change in NHAA brains.

To determine whether neuropathology associated with metabolites differently across ethnoracial groups, each metabolite was examined with a model that included Braak stage, ethnoracial group, and their interaction. This approach identified 60/907 metabolites (6.6%) levels of which showed a significant or suggestive Braak-by-ethnoracial interaction after FDR adjustment (p < 0.1). By contrast, the same analysis based on CERAD score, Reagan score, and AD diagnosis revealed no significant interactions (see **Supplementary Data 2**). The class distribution of the 60 Braak-related interaction hits resembled the background distribution, with lipids again representing the predominant group (41.7%). In addition, energy metabolites, consisting of tricarboxylic-acid-cycle intermediates, and dipeptides were significantly over-represented relative to their baseline frequencies (Fisher’s exact test, p = 0.006 and p = 0.042, respectively, **Figure 3a, right**).

Acetylcholine, the only neurotransmitter among the 60 significant metabolites, exhibited significant ethnoracial heterogeneity (FDR-adjusted p < 0.1), with all three groups showing a decline in abundance with advancing Braak stage. However, the magnitude of decline differed markedly, with a much more pronounced effect in HAs as compared to NHWs and NHAAs (**Supplementary Figures**).

Fumarate, malate, and succinate showed divergent behavior by ethnoracial group with NHAAs differing in metabolite trajectories as compared to HA and NHW groups (**Figure 3b**). Interaction testing confirmed significant ethnoracial heterogeneity for all three TCA-cycle metabolites, but the source of this heterogeneity was metabolite-specific (**Supplementary Data 3**). Follow-up, within-group-specific models showed that for fumarate, the interaction signal was driven primarily by a significant positive association in HAs, with no detectable relationship in NHWs or NHAAs; for succinate, robust associations were present in both HAs and NHWs but absent in NHAAs; and for malate, the heterogeneity arose from a weak, non-significant trend in HAs, with no association in the other groups.

Eight dipeptides met the interaction significance threshold (**Figure 3c** and **Supplementary Figures)**. In Hispanic American donors, dipeptide abundance rose sharply with advancing Braak stage while Non-Hispanic Whites showed a more modest upward trend and non-Hispanic African Americans displayed no detectable change. This was confirmed by within-group analysis, where only the HA group shows consistent significant associations between Braak and the dipeptides (**Supplementary Data 3**).

### Selective reduction of sphingomyelins and ceramides distinguishes Braak stage progression in ethnoracial groups

Examination of the strongest signals revealed a coherent lipid-centric theme: ten of the fifteen most significant hits (66.7%) mapped to the ceramide/sphingomyelin (SM) pathway (**Table 2**, **Figure 4a**). As Braak stage increased, SM and ceramide species declined in both NHW and HA donors, whereas levels remained unchanged in NHAAs. Within-group analysis confirmed that there were significant associations between Braak stage and these lipids specifically in the HA and NHW groups (**Supplementary Data 3**). This collective downward trajectory was consistent across all heterogeneous metabolites in the pathway, indicating a coordinated rewiring of sphingolipid metabolism with advancing tau pathology in NHWs and HAs, but not NHAAs (**Figure 4b**).

**Figure 4:**
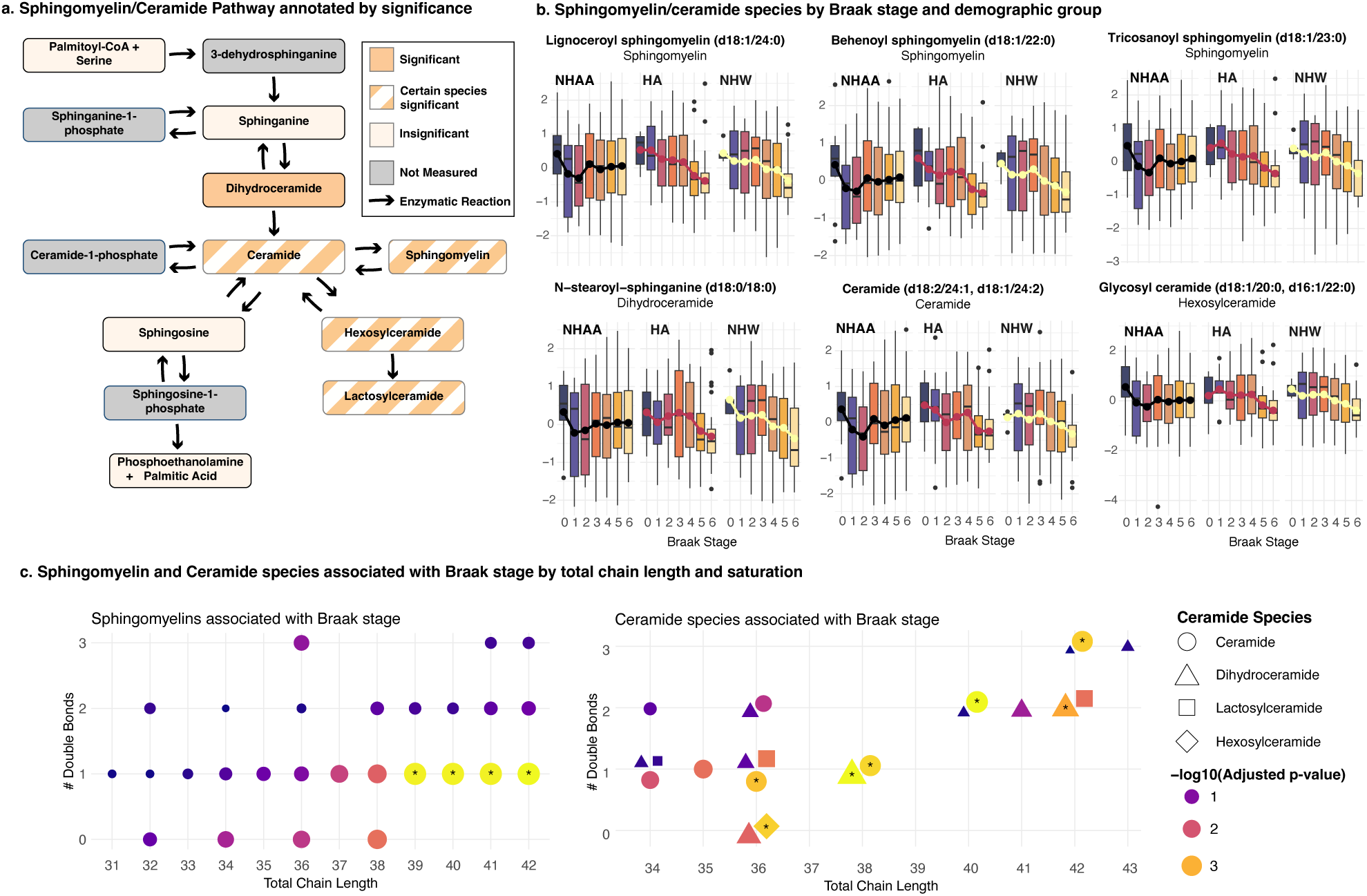
**a** Ceramide/Sphingomyelin Pathway. Adapted from Baloni et al, 2022^47^. Divergent metabolites were enriched in this pathway. **b** Sphingomyelin and ceramide species by Braak stage and demographic group. Across the species of sphingomyelins and ceramides that show significant divergence between demographic groups, the same pattern emerges: as Braak stage progresses, there is a decrease in the metabolites’ abundance amongst Hispanic American and White non-Hispanic subjects, while there is no change in African American non-Hispanic subjects. This patter persists for all sphingomyelins and ceramides identified. **c** Significant divergence in ceramides/sphingomyelins by chain length and saturation. Color represents –log10(nominal p-value), stars indicate significant divergent hits. While the ceramide/sphingomyelin pathway was enriched among the divergent hits, not all species showed heterogeneous behavior by demographic group. Of the sphingomyelins with and 18:1 sphingosine chain, those with fatty acid chains with over 20 carbons showed divergent behavior.

**Table 2.** 15 most significant metabolite interactions with ethnoracial group.

| Metabolite | Outcome | Adjusted p-value | Molecular Classification |
| --- | --- | --- | --- |
| N-carboxymethylalanine | Braak | 0.0133 | Food Component/Plant |
| Lignoceroyl sphingomyelin (d18:1/24:0) | Braak | 0.0212 | Sphingomyelins |
| Behenoyl sphingomyelin (d18:1/22:0) | Braak | 0.0227 | Sphingomyelins |
| Sphingomyelin (d18:1/21:0, d17:1/22:0, d16:1/23:0) | Braak | 0.0229 | Sphingomyelins |
| Tricosanoyl sphingomyelin (d18:1/23:0) | Braak | 0.0229 | Sphingomyelins |
| Glycosyl ceramide (d18:1/20:0, d16:1/22:0) | Braak | 0.0240 | Hexosylceramides (HCER) |
| N-behenoyl-sphingadienine (d18:2/22:0) | Braak | 0.0240 | Ceramides |
| Pantothenate | Braak | 0.0240 | Pantothenate and CoA Metabolism |
| 1-stearoyl-2-docosapentaenoyl-GPS (18:0/22:5n3) | Braak | 0.0335 | Phosphatidylserine (PS) |
| Ceramide (d18:1/20:0, d16:1/22:0, d20:1/18:0) | Braak | 0.0335 | Ceramides |
| Ceramide (d18:2/24:1, d18:1/24:2) | Braak | 0.0335 | Ceramides |
| Maltotetraose | Braak | 0.0335 | Glycogen Metabolism |
| N-stearoyl-sphinganine (d18:0/18:0) | Braak | 0.0335 | Dihydroceramides |
| Kynurenate | Braak | 0.0359 | Tryptophan Metabolism |
| N-stearoyl-sphingosine (d18:1/18:0) | Braak | 0.0359 | Ceramides |

To pinpoint the biochemical features driving this signal, we stratified sphingomyelins and ceramides by fatty-acyl chain length and degree of saturation (**Figure 4c**). Among SMs, only species with very-long-chain fatty acids (> C20) showed significant interactions between Braak stage and ethnoracial group. Shorter-chain SMs (< C20) did not show comparable divergence. Thus, the ethnoracial heterogeneity seems to be driven by selective tau-dependent loss of very-long-chain SMs in NHW and Hispanic American brains. In contrast, ceramides showed interaction signals at a handful of chain-length/saturation combinations, but these were not enriched for particular biochemical properties.

Together with our findings related to dipeptide- and energy-pathways, these results reveal a multi-layered, population-based metabolic heterogeneity in AD that spans proteolysis, bioenergetics, and, most prominently, sphingolipid remodeling.

## Discussion

In this study, we applied untargeted metabolomics to a large, multiethnic brain cohort, including dorsolateral prefrontal cortex tissue from 545 postmortem donors from 220 non-Hispanic African Americans, 148 Hispanic Americans and 177 non-Hispanic Whites. As these samples originated from four different study sites, stringent quality control and batch correction was required to reduce site attributable variance from ∼4 % to <0.5 % per metabolite while preserving biological signals. By modelling each of 907 metabolites as a function of four neuropathology measures of AD, and their interaction with ethnoracial group membership, we found that only Braak stage, which reflects neurofibrillary-tangle pathology, interacted with ethnoracial group. Reagan score, CERAD score, and clinical diagnosis showed no significant interaction with ethnoracial group. This is consistent with our previous work in NHWs, where we identified tau load as a potential driver of metabolic dysfunction in the late-stage AD brain, with minimal contributions from Aβ load^20^. We speculate that tau’s prominent role in driving metabolic dysfunction in the postmortem brain may underlie these group-specific differences emerging only for the tau-related neuropathological measures. In total, sixty metabolites showed significant Braak-by-group interactions, revealing divergent trajectories of the neurotransmitter acetylcholine, driven by a sharper decline in Hispanic Americans as compared to shallower declines in other groups; key tricarboxylic acid cycle intermediates, driven mainly by Hispanic American samples; a steep Braak-dependent rise in dipeptides unique to Hispanic Americans; and a pathway-wide, selective depletion of very-long-chain sphingomyelins (with scattered ceramide changes) in White and Hispanic American brains but not in African Americans.

The population-level heterogeneity in acetylcholine decline could have implications for drug efficacy and dosing of cholinesterase inhibitors, a class of AD treatments that act by increasing acetylcholine availability^39^. The differences uncovered here for the first time in brain tissue, suggest that response to cholinesterase-based therapy may vary by ethnoracial group and should be considered in both clinical trial design and treatment planning.

Overall, our findings indicate that tau pathology engages distinct lipid, peptide-catabolic and bioenergetic programs according to ethnoracial background, underscoring the need for population-aware mechanistic models and therapeutic strategies in Alzheimer’s disease.

### Sphingomyelin/ceramide pathway exhibits population-specific disruption

The present findings revealed a tau-linked disruption of sphingolipid metabolism that appears to differ by ethnoracial group. This is consistent with previous studies that have already reported correlations between myelin levels, metabolites, or transcriptome with tau pathology^40–46^. In our data we observed that in non-Hispanic White (NHW) and Hispanic American (HA) brains, advancing tau pathology (indicated by higher Braak stage) was accompanied by a selective loss of long-chain sphingomyelins (SM) and a parallel decline in ceramides. Only SM species with fatty acyl chains >C20, which are highly enriched in myelin sheaths^47,48^, showed this tau-associated reduction^43^. These results indicate that that during late-stage tau pathology, sphingolipid pools might become exhausted, or ceramides might be further metabolized, resulting in net lower levels. Regardless of the underlying mechanism, the tau-associated loss of long-chain SMs in NHW and Hispanics points to active myelin degradation and lipid turnover, a process that could contribute to neurodegeneration and is consistent with known white-matter damage in AD.

The absence of these sphingolipid changes in non-Hispanic African Americans (NHAAs) underscores how population diversity can unveil non-universal disease patterns. In our NHAA group, SM and ceramide levels remained stable across Braak stages, indicating a relative preservation of sphingolipid homeostasis despite accumulating tau. While this divergence within brain tissue is a novel finding, prior research hints at similar ethnoracial differences in fluids. For example, a plasma metabolomics study found that AD NHWs and Caribbean Hispanics showed altered glycosphingolipid metabolism, whereas African American patients did not exhibit such lipid perturbations^49^. Additionally, African Americans have been reported to display lower brain tau biomarker levels in CSF as compared to whites with the same degree of cognitive impairment^50^. Such findings suggest that tau-associated myelin damage or lipid release might be less pronounced in NHAAs potentially due to genetic, environmental, or comorbidity factors that modulate disease pathophysiology. This interpretation is further supported by evidence that the allelic dosage of *APOE4*, which plays a central role in brain lipid metabolism, results in slower decline in cognitive tests in NHAAs as compared to NHWs^12^, and less disease risk in individuals of African ancestry as compared to European ancestry^51^.

Our results therefore emphasize that including diverse participants can reveal divergent biochemical responses to AD pathology that would be overlooked in homogeneous subjects. This has significant clinical implications: in practice, a candidate therapy aimed at bolstering myelin lipids^52^ or normalizing ceramide levels might show varying efficacy across populations unless these differences are understood. Overall, the tau–sphingolipid findings in this study highlight a multi-layered, population-dependent heterogeneity in AD.

### Divergent dipeptide and TCA trajectories suggest heterogeneous proteostatic and bioenergetic stress

Beyond sphingolipid remodeling, we observed population-specific shifts in proteolytic and bioenergetic pathways associated with tau pathology. Products of protein degradation in the form of dipeptides, were disproportionately represented among metabolites showing Braak-stage interactions, with levels rising sharply in Hispanic Americans, increasing modestly in non-Hispanic Whites (NHWs), and remaining stable in non-Hispanic African Americans (NHAAs). While the connection between dipeptides and AD is not clear, some evidence shows that the dipeptide carnosine, normally abundant in brain tissue and cerebrospinal fluid, is depleted in AD and has been linked to impaired cognitive function and reduced cerebral vasodilation^53,54^. Loss of carnosine and other dipeptides with antioxidant or vasoregulatory roles may exacerbate neurovascular dysfunction in AD, a mechanism potentially more active in Hispanic American brains where dipeptide accumulation may reflect compensatory or pathological proteostatic stress. In support of this, studies in hypertensive rat models show that certain orally delivered dipeptides can reverse neuronal damage and cognitive deficits by activating pro-survival signaling and reducing apoptosis^55^, further underscoring the potential vascular and metabolic significance of dipeptide regulation in AD.

Energy metabolites, specifically TCA-cycle intermediates fumarate, malate, and succinate, also showed divergent trajectories across ethnoracial groups. In NHWs and HAs, levels of these metabolites shifted with increasing Braak stage, whereas NHAAs showed distinct or flat patterns. These findings align with well-established mitochondrial dysfunction in AD brain tissue, where impaired TCA cycling and oxidative phosphorylation lead to energy deficits, oxidative stress, and neurodegeneration^56–60^. Mechanistically, fumarate, malate, and succinate lie downstream of succinyl-CoA. Common variation in SUCLG2, the GDP-specific β subunit of mitochondrial succinyl-CoA synthetase, has been associated with AD phenotypes and cognitive decline^61^. While mitochondrial failure is a common feature of AD, emerging evidence suggests it may unfold differently across populations. For example, oxidative damage to mitochondrial DNA has been reported to be elevated in Mexican American AD patients compared to NHWs, correlating with worse cognitive performance^62^. Mitochondrial haplogroup associations with AD risk have also been explored, suggesting that ancestry-related mtDNA variants may modulate susceptibility^63^. The absence of consistent TCA metabolite shifts in NHAAs could indicate more stable mitochondrial function in this group during tau accumulation, or it may reflect distinct mechanisms of metabolic adaptation or resilience.

These population-specific patterns in proteolysis and mitochondrial metabolites reinforce the idea that AD is not a monolithic disease. Pathophysiological responses, including how the brain processes damaged proteins and maintains energetic homeostasis, may differ by ancestry due to genetic, environmental, or vascular co-factors. Without samples from multi-ethnic cohorts representing different ancestries and variable exposures, such divergent trajectories would remain obscured, potentially leading to incomplete or biased models of AD biology. Understanding these differences is not only critical for interpreting disease mechanisms, but also for identifying biomarkers and tailoring interventions that reflect the full spectrum of disease across populations.

### Complexities of self-reported ethnoracial grouping

Our analysis rests on three self-identified categories: non-Hispanic African American, Hispanic American, and non-Hispanic White. These categories sit at the intersection of genetic ancestry and culture. They capture broad continental ancestries but also likely embed social parameters such as residential segregation, differential access to care, diet, education, and chronic stress. These socially structured exposures have been increasingly linked to molecular phenotypes, including emerging evidence that they shape the metabolome^64–66^. Within-group heterogeneity is therefore substantial: a “Latin/Hispanic” donor may carry Indigenous, African, and European genetic traits in proportions that vary by birthplace and family history, while an “African American” donor from the U.S. Southeast may differ markedly in both genome and lived experience from one raised in California.

Relying solely on genetic ancestry for grouping, while common in research, risks overlooking important sociocultural and environmental determinants of biology and health. The limitations of purely genetic definitions are underscored by recent advances in genomics, demonstrating that our existing genomic references, and by extension ancestry-based frameworks built upon them, are incomplete and biased against underrepresented populations^67^. Likewise, polygenic risk score analyses across diverse African populations have shown that prediction accuracy varies widely not only between continental ancestries but also within Africa, where differences in local ancestry can produce variability in predictive power as large as those between continents^68^. These findings highlight that genetic ancestry alone cannot fully capture the biological, cultural, and environmental complexity influencing disease risk or metabolism. Consequently, the metabolite-by-group interactions we report cannot be assigned unambiguously to germ-line genetics, social environment, or their interaction.

This ambiguity has two implications. First, the broad ethnoracial labels themselves may dilute true effects: because each self-identified category spans a wide spectrum of genetic ancestries and social experiences, the metabolic signals we detect are probably weakened. Second, mechanistic inference is clouded: for example, selective loss of long-chain sphingomyelins in HAs and NHWs could reflect allele-frequency differences in lipid-metabolism genes, but it could equally index diet, metabolic comorbidity, or medication patterns that covary with those labels.

In future work, coupling genome-wide ancestry proportions with granular social and lifestyle metrics, including education, neighborhood, diet, and stress burden, will be essential for untangling social from biological drivers and for ensuring that metabolic biomarkers generalize across the full spectrum of human diversity.

### Limitations and caveats

Several factors limit the generalizability of our findings.

First, residual site effects and pre-analytical variability may affect ethnoracial associations. We combined tissue from four U.S. brain banks that differed in autopsy workflow, storage logistics and donor composition. Although ComBat reduced the fraction of variance attributable to “site” from ∼4 % to <0.5 % per metabolite, the lack of NHWs at Mount Sinai and of HAs at Rush means that ancestry and site remain partially confounded. Moreover, variations in parameters known to influence brain metabolite read-outs, including freezing protocols or storage temperature^69,70^, could leave artefacts that evade correction. Standardized, prospective tissue-collection pipelines in future studies will be essential to confirm the ancestry-linked sphingolipid signal we report.

Second, our study lacked complete clinical and lifestyle metadata. Key modifiers of brain metabolism, including years of education, body-mass index, diet and physical activity were unavailable for most donors, and detailed medication histories were lacking as well. Drugs that alter lipid or energy metabolism (e.g., statins, antihypertensives, GLP-1 agonists) might therefore confound some of the group-specific trajectories we observe. Future studies that integrate richer covariate sets will be required to disentangle social, behavioral and therapeutic influences from ethnic and racial groupings.

Third, missing neuropathological endpoints limited our statistical power. Our ancestry-by-Braak interaction screen benefited from the largest complete-case sample (n = 545), but the CERAD, Reagan and AD-diagnosis subsets were roughly 20% smaller and may have been under-powered to detect comparable heterogeneity. The absence of interaction hits for those endpoints should therefore be interpreted with caution rather than as clear evidence of pathologic specificity.

Fourth, all measurements derive from a single cortical region (dorsolateral prefrontal cortex, DLPFC) at one postmortem time-point. There may be brain-region specific metabolite changes. For example, we previously discovered that ceramides are upregulated in the superior temporal gyrus of AD brains^44^, in contrast to the DLPFC findings here. Multi-region brain metabolome profiling as well as longitudinal sampling of blood or CSF from antemortem cohorts would be needed to establish whether the ethnoracial-linked signatures we describe have brain regional specificity and precede, accompany or result from tau spread.

Finally, bulk-tissue metabolomics lacks the resolution to identify cell-type-specific contributions to the observed associations. Differences in the relative abundance of neurons, glia, and other cell types as well as cell-type specific metabolic reprogramming in response to pathology could alter the bulk metabolic profile, convoluting downstream interpretation^71–73^. Future work using single-cell or spatially resolved metabolomics is needed to determine whether the heterogeneity we observe is rooted in cell composition, cell-specific metabolic changes, or both.

### Conclusion and future directions

By harmonizing untargeted metabolomic profiles from more than five hundred postmortem brains across four U.S. sites, we assembled the largest ethnoracially diverse brain metabolomics resource to date. The dataset captures how self-identified ancestry, and its intertwined social milieus, correlate with Alzheimer’s disease biochemistry. This included ethnoracial-specific shifts in sphingolipid remodeling, peptide catabolism, and mitochondrial fuel use. These findings provide a first step towards a molecular rationale for tailoring both biomarkers and therapies to the populations they are meant to serve. For example, our data suggest that long-chain sphingomyelin loss is muted in African Americans; therapeutic window may therefore differ between ethnoracial groups, an important consideration for more efficacious clinical trials. More broadly, this work offers a foundation for dissecting how demographic variables intersect with distinct metabolic pathways, neuropathologic markers, and druggable targets in Alzheimer’s disease, accelerating progress toward truly precise, population-specific interventions.

## Acknowledgements

The results published here are in whole or in part based on data obtained from the AD Knowledge Portal Diverse Cohort Study (https://doi.org/10.7303/9618093). Data generation was supported by the following NIH grants: U01AG046139, U01AG046170, U01AG061357, U01AG061356, U01AG061359, and R01AG067025. We thank the participants of participants of the Religious Order Study, Memory and Aging Project, the Minority Aging Research Study, Rush Alzheimer’s Disease Research Center, Mount Sinai/JJ Peters VA Medical Center NIH Brain and Tissue Repository, National Institute of Mental Health Human Brain Collection Core (NIMH HBCC), Mayo Clinic Brain Bank, Sun Health Research Institute Brain and Body Donation Program, Goizueta Alzheimer’s Disease Research Center, New York Brain Bank at Columbia University, New York Genome Center and the Biggs Institute Brain Bank for their generous donations. Data and analysis contributing investigators include Nilüfer Ertekin-Taner, Minerva Carrasquillo, Mariet Allen (Mayo Clinic, Jacksonville, FL), David Bennett, Lisa Barnes (Rush University), Philip De Jager, Vilas Menon (Columbia University), Bin Zhang, Vahram Haroutanian (Icahn School of Medicine at Mount Sinai), Allan Levey, Nick Seyfried (Emory University), Rima Kaddurah-Daouk (Duke University), Steve Finkbeiner (University of California-San Francisco/Gladstone Institutes), Daifeng Wang (University of Wisconsin-Madison), Stefano Marenco (NIMH HBCC), Anna Greenwood, Abby Vander Linden, Laura Heath, William Poehlman (Sage Bionetworks).

## Data Availability

Data used in this manuscript are available via the AD Knowledge Portal (https://adknowledgeportal.org). The AD Knowledge Portal is a platform for accessing data, analyses, and tools generated by the Accelerating Medicines Partnership (AMP-AD) Target Discovery Program and other National Institute on Aging (NIA)-supported programs to enable open-science practices and accelerate translational learning. The data, analyses and tools are shared early in the research cycle without a publication embargo on secondary use. Data is available for general research use according to the following requirements for data access and data attribution (https://adknowledgeportal.synapse.org/Data%20Access). For access to content described in this manuscript see: https://doi.org/10.7303/9618093.

## Supplement Captions

Supplementary Material 1: Metabolon Methods

Supplementary Table 1: Descriptive characteristics of 716 dorsolateral prefrontal cortex samples before filtration due to missing covariate information

Supplementary Table 2: Distributions of neuropathology measures across ethnoracial groups

Supplementary Data 1: Molecular annotations of 1388 metabolites collected in the current analysis. All annotations provided by the Metabolon platform

Supplementary Data 2: Results of MWAS interaction analysis of 907 metabolites across 4 Alzheimer’s disease neuropathological measures (CERAD score, Braak Stage, NIA Reagan score, and NP diagnosis)

Supplementary Data 3: Metabolites with significant interaction with AD neuropathology by ethnoracial group (FDR p-value < 0.1)

Supplementary Figures: All metabolites with a significant interaction with Braak stage by ethnoracial group (FDR p-value < 0.1). Asterisks represent within-group significance results. ***: p < 0.005, **: p < 0.01, *: p < 0.05.

