## Supplementary Table 1 for "Sphingolipid and ceramide associations with tau pathology vary across diverse ethnoracial groups in postmortem brain tissue"

Supplementary Table 1: Descriptive characteristics of 716 dorsolateral prefrontal cortex samples before filtration due to missing covariate information

| **Characteristic** | **Overall**  N = 716*^1^* | **Emory**  N = 80*^1^* | **Mayo Clinic**  N = 219*^1^* | **Mt Sinai Brain Bank**  N = 56*^1^* | **RADC**  N = 361*^1^* |
| --- | --- | --- | --- | --- | --- |
| Sex |  |  |  |  |  |
| Female | 454 (64%) | 47 (59%) | 114 (52%) | 32 (57%) | 261 (74%) |
| Male | 256 (36%) | 33 (41%) | 105 (48%) | 24 (43%) | 94 (26%) |
| Unknown | 6 | 0 | 0 | 0 | 6 |
| Number of APOE4 alleles |  |  |  |  |  |
| 0 | 395 (60%) | 31 (40%) | 126 (58%) | 33 (60%) | 205 (67%) |
| 1 | 213 (32%) | 35 (45%) | 78 (36%) | 20 (36%) | 80 (26%) |
| 2 | 50 (7.6%) | 12 (15%) | 15 (6.8%) | 2 (3.6%) | 21 (6.9%) |
| Unknown | 58 | 2 | 0 | 1 | 55 |
| Alzheimer's Disease Diagnosis |  |  |  |  |  |
| Case | 424 (60%) | 61 (76%) | 126 (58%) | 38 (68%) | 199 (56%) |
| Control | 119 (17%) | 18 (23%) | 14 (6.4%) | 11 (20%) | 76 (21%) |
| Other*^2^* | 167 (24%) | 1 (1.3%) | 79 (36%) | 7 (13%) | 80 (23%) |
| Unknown | 6 | 0 | 0 | 0 | 6 |
| Post-mortem Interval (hours) | 8 (6, 14) | 7 (5, 15) | 12 (5, 21) | 7 (4, 10) | 8 (6, 12) |
| Unknown | 105 | 0 | 99 | 0 | 6 |
| Age at death (years) | 84 (76, 90) | 75 (61, 82) | 79 (71, 85) | 82 (75, 90) | 89 (82, 93) |
| Unknown | 7 | 0 | 1 | 0 | 6 |
| Ethnoracial Group |  |  |  |  |  |
| African American non-Hispanic | 255 (36%) | 42 (53%) | 47 (21%) | 28 (50%) | 138 (38%) |
| Hispanic American | 248 (35%) | 0 (0%) | 147 (67%) | 27 (48%) | 74 (20%) |
| White non-Hispanic | 213 (30%) | 38 (48%) | 25 (11%) | 1 (1.8%) | 149 (41%) |
| *^1^*n (%); Median (Q1, Q3) | | | | |  |
| *^2^*Did not meet case or control criteria (Case: Braak stage ≥ 4 and CERAD score ≤ 2; Control: Braak stage ≤ 3 and CERAD ≥ 3) | | | | |  |
