## Supplementary Table 2 for "Sphingolipid and ceramide associations with tau pathology vary across diverse ethnoracial groups in postmortem brain tissue"

Supplementary Table 2: Distributions of neuropathology measures across ethnoracial groups

| **Characteristic** | **Hispanic American**  N = 148*^1^* | **Non-Hispanic African American**  N = 222*^1^* | **Non-Hispanic White**  N = 177*^1^* |
| --- | --- | --- | --- |
| Study Site |  |  |  |
| Emory | 0 (0%) | 40 (18%) | 37 (21%) |
| Mayo Clinic | 73 (49%) | 31 (14%) | 15 (8.5%) |
| Mt Sinai Brain Bank | 27 (18%) | 28 (13%) | 0 (0%) |
| RADC | 48 (32%) | 123 (55%) | 125 (71%) |
| Reagan Score |  |  |  |
| 1 | 28 (37%) | 76 (40%) | 51 (31%) |
| 2 | 27 (36%) | 52 (27%) | 56 (35%) |
| 3 | 17 (23%) | 58 (30%) | 51 (31%) |
| 4 | 3 (4.0%) | 5 (2.6%) | 4 (2.5%) |
| Unknown | 73 | 31 | 15 |
| CERAD Score |  |  |  |
| 0 | 14 (19%) | 44 (23%) | 38 (23%) |
| 1 | 4 (5.3%) | 17 (8.9%) | 13 (8.0%) |
| 2 | 21 (28%) | 31 (16%) | 48 (30%) |
| 3 | 36 (48%) | 99 (52%) | 63 (39%) |
| Unknown | 73 | 31 | 15 |
| Braak Stage |  |  |  |
| 0 | 6 (4.1%) | 10 (4.5%) | 5 (2.8%) |
| 1 | 11 (7.4%) | 12 (5.5%) | 11 (6.2%) |
| 2 | 16 (11%) | 21 (9.5%) | 15 (8.5%) |
| 3 | 18 (12%) | 34 (15%) | 28 (16%) |
| 4 | 28 (19%) | 48 (22%) | 45 (25%) |
| 5 | 27 (18%) | 51 (23%) | 48 (27%) |
| 6 | 42 (28%) | 44 (20%) | 25 (14%) |
| Unknown | 0 | 2 | 0 |
| Alzheimer's Disease Diagnosis |  |  |  |
| Case | 90 (61%) | 127 (57%) | 105 (59%) |
| Control | 14 (9.5%) | 46 (21%) | 44 (25%) |
| Other*^2^* | 44 (30%) | 49 (22%) | 28 (16%) |
| *^1^*n (%) | | | |
| *^2^*Did not meet case or control criteria (Case: Braak stage ≥ 4 and CERAD score ≤ 2; Control: Braak stage ≤ 3 and CERAD ≥ 3) | | | |
