## Supplementary figures and images for "Sphingolipid and ceramide associations with tau pathology vary across diverse ethnoracial groups in postmortem brain tissue"

### Supplementary Figures

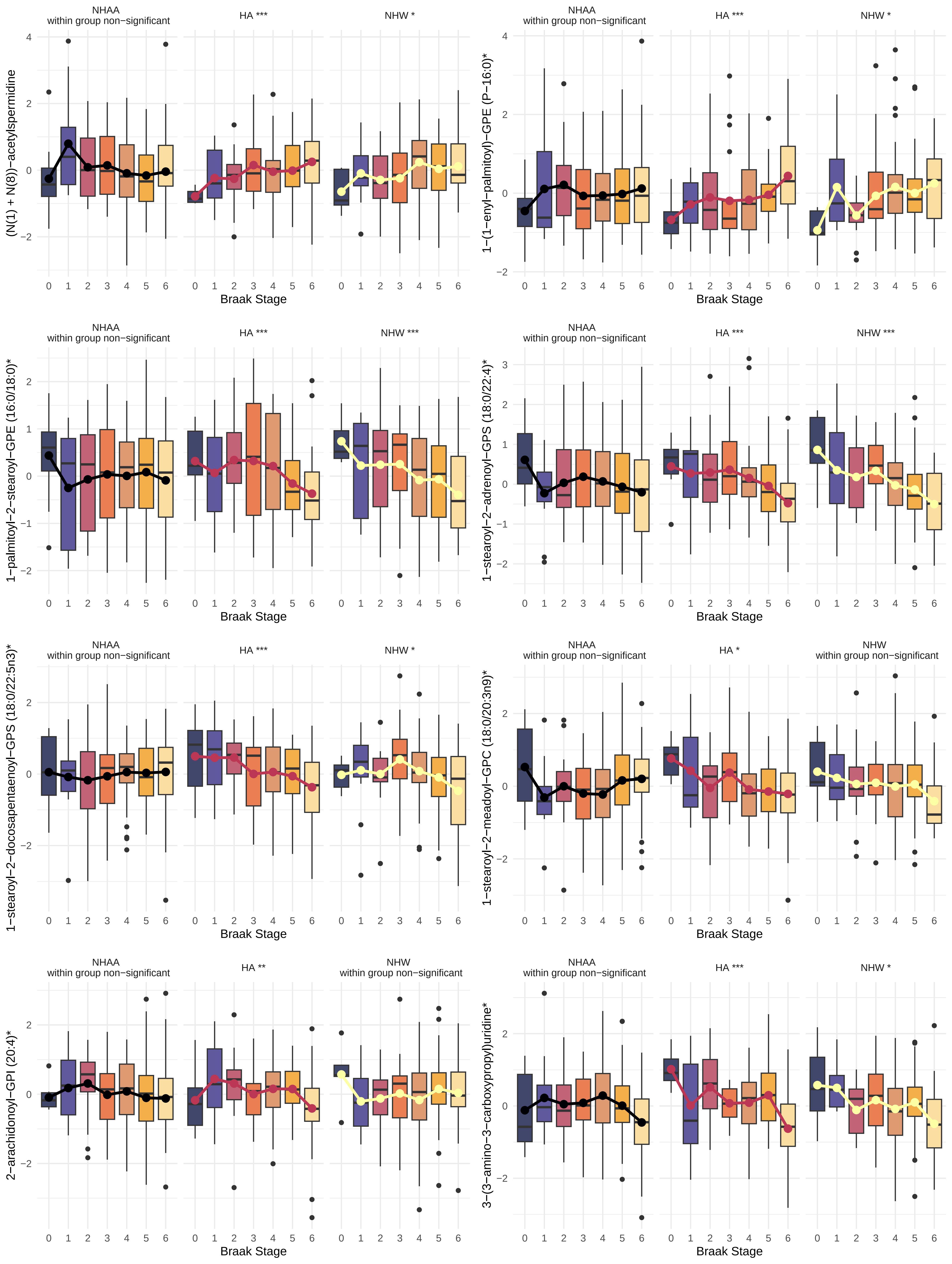

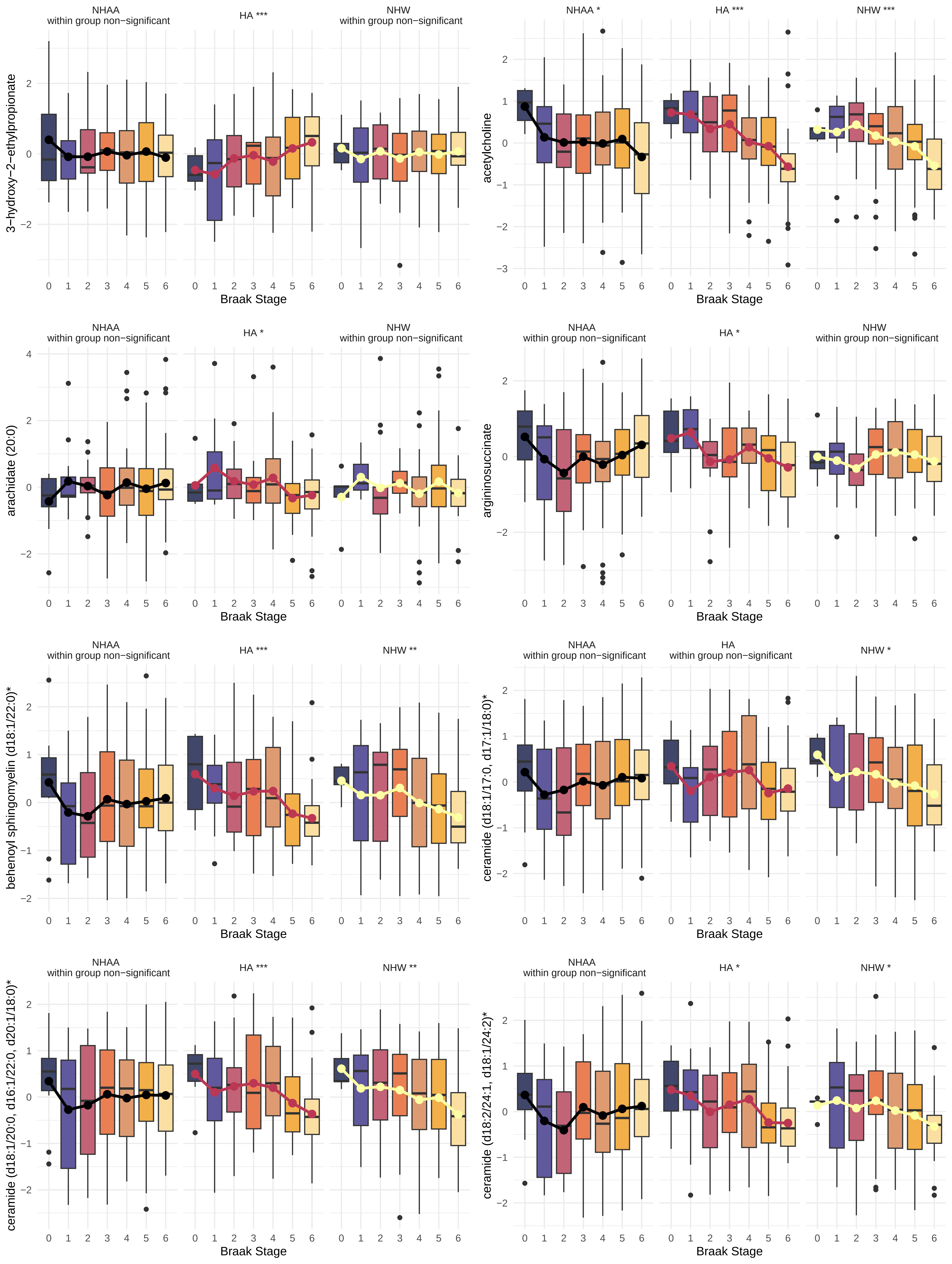

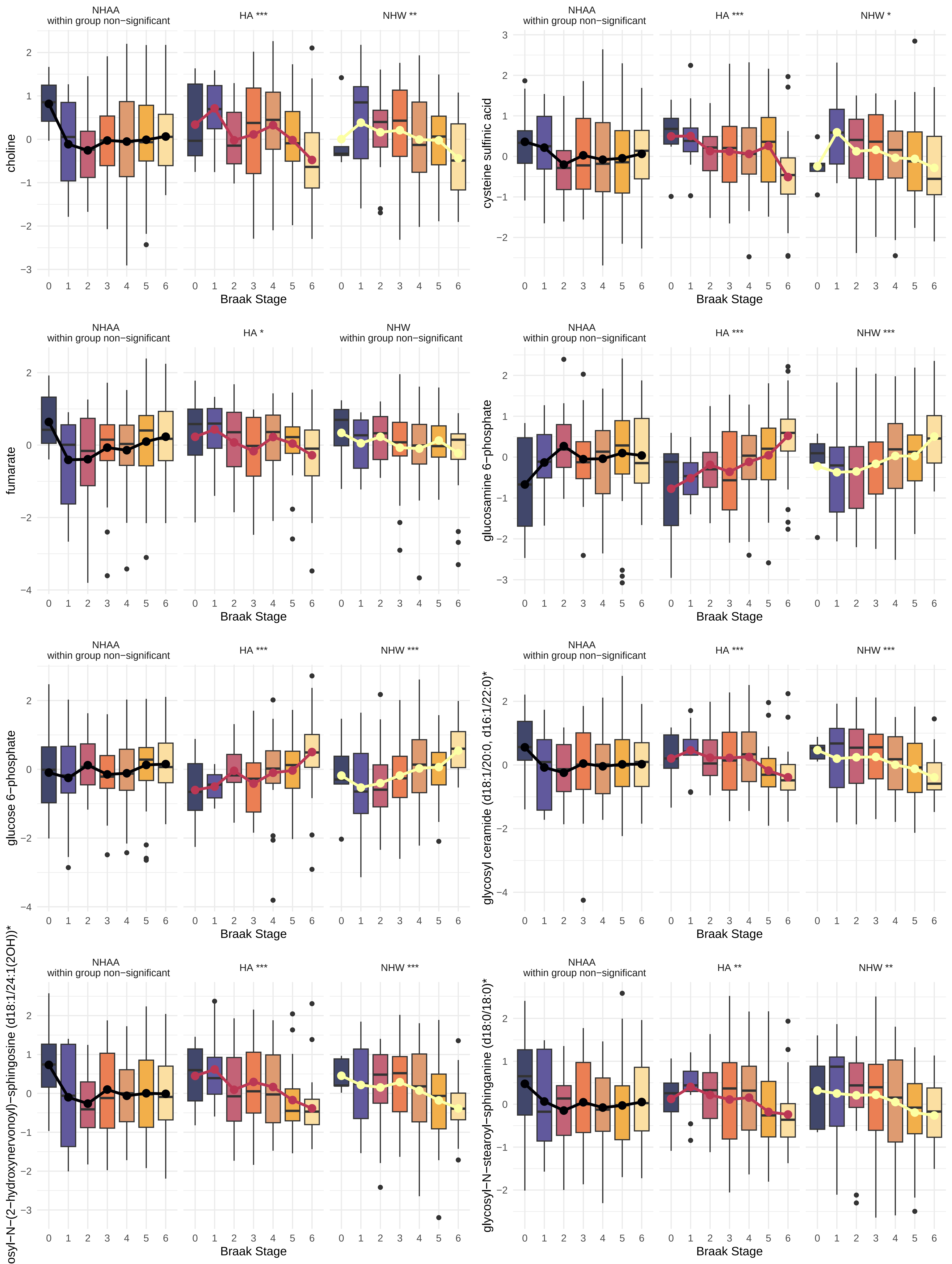

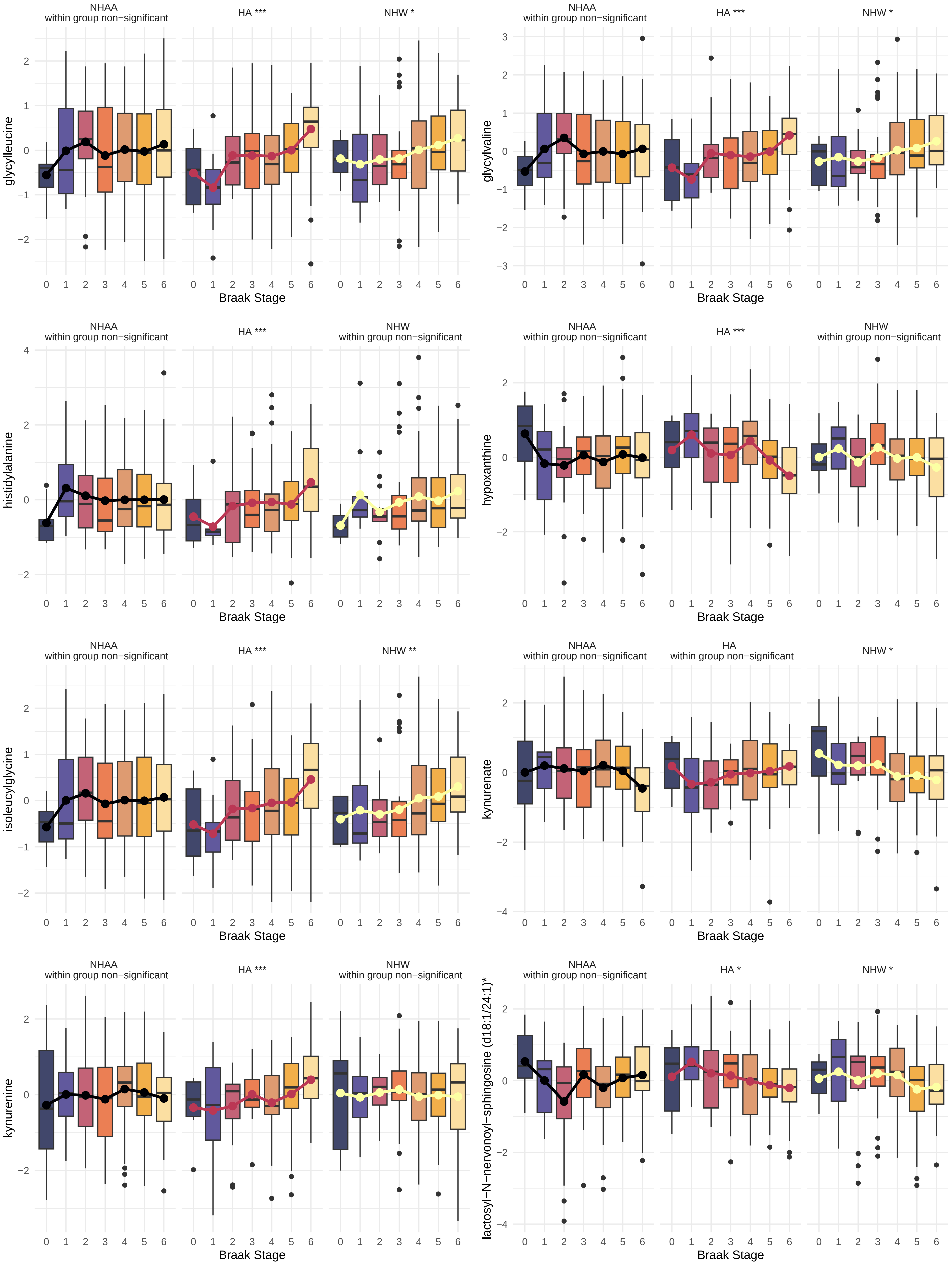

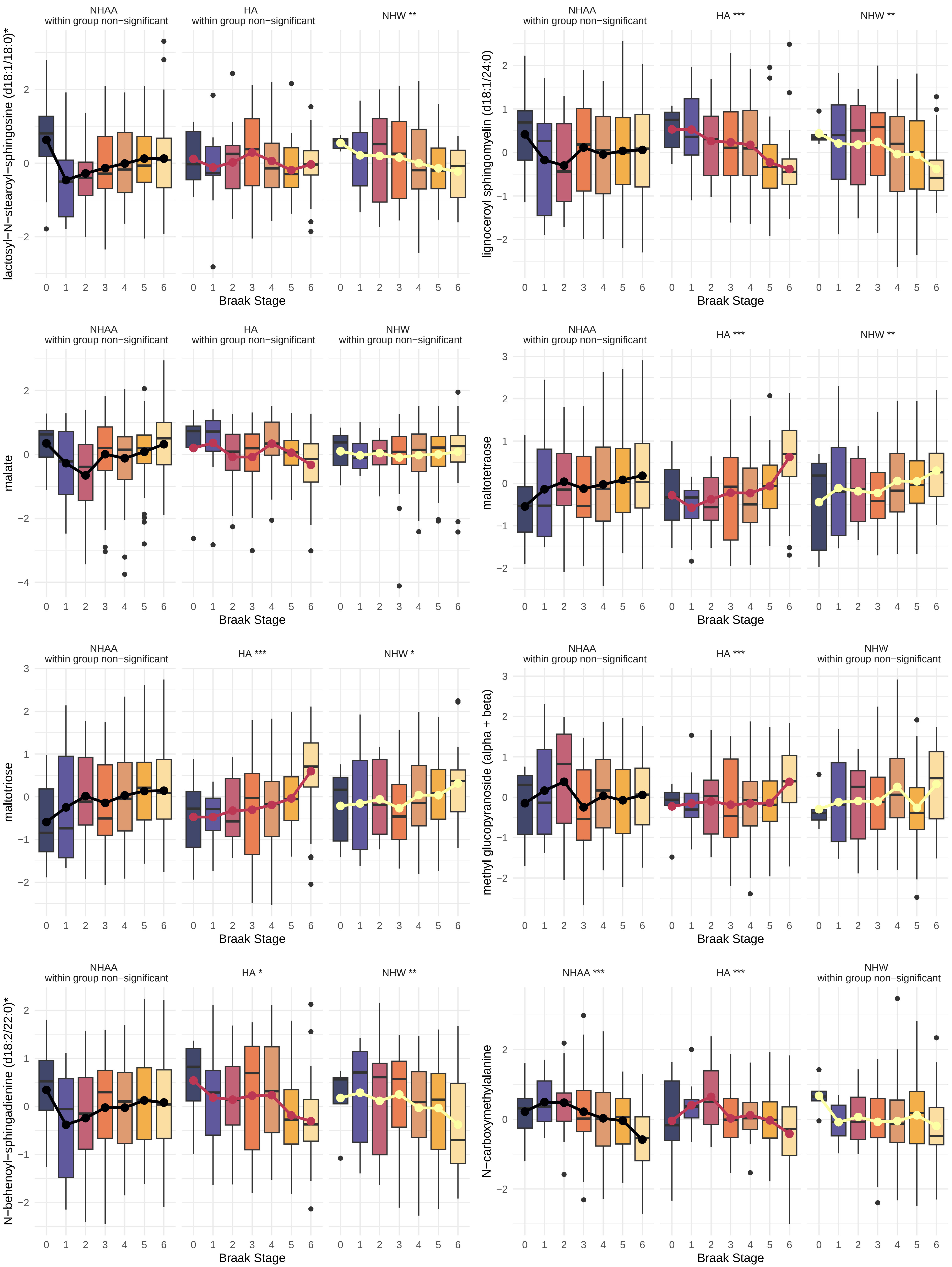

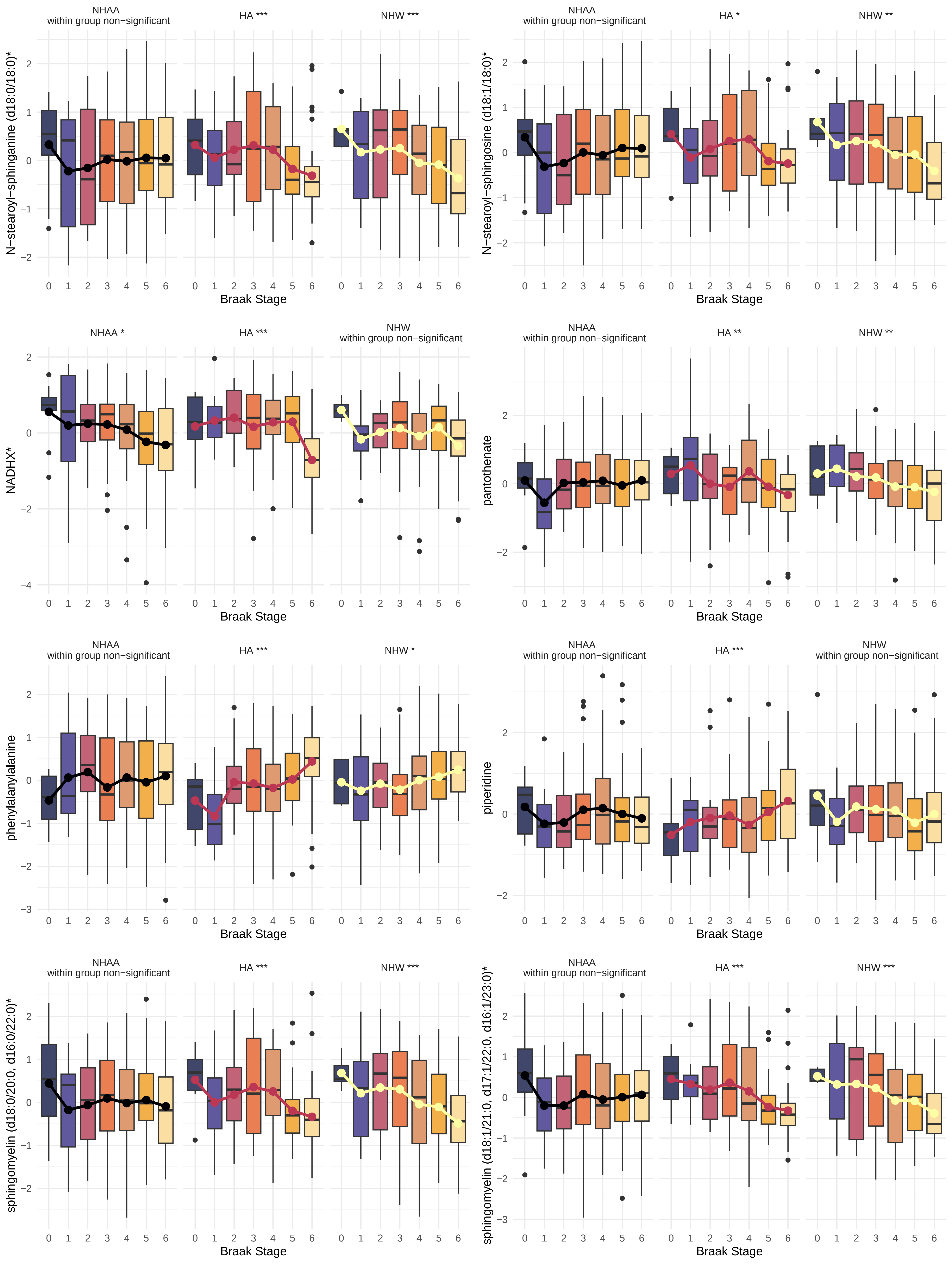

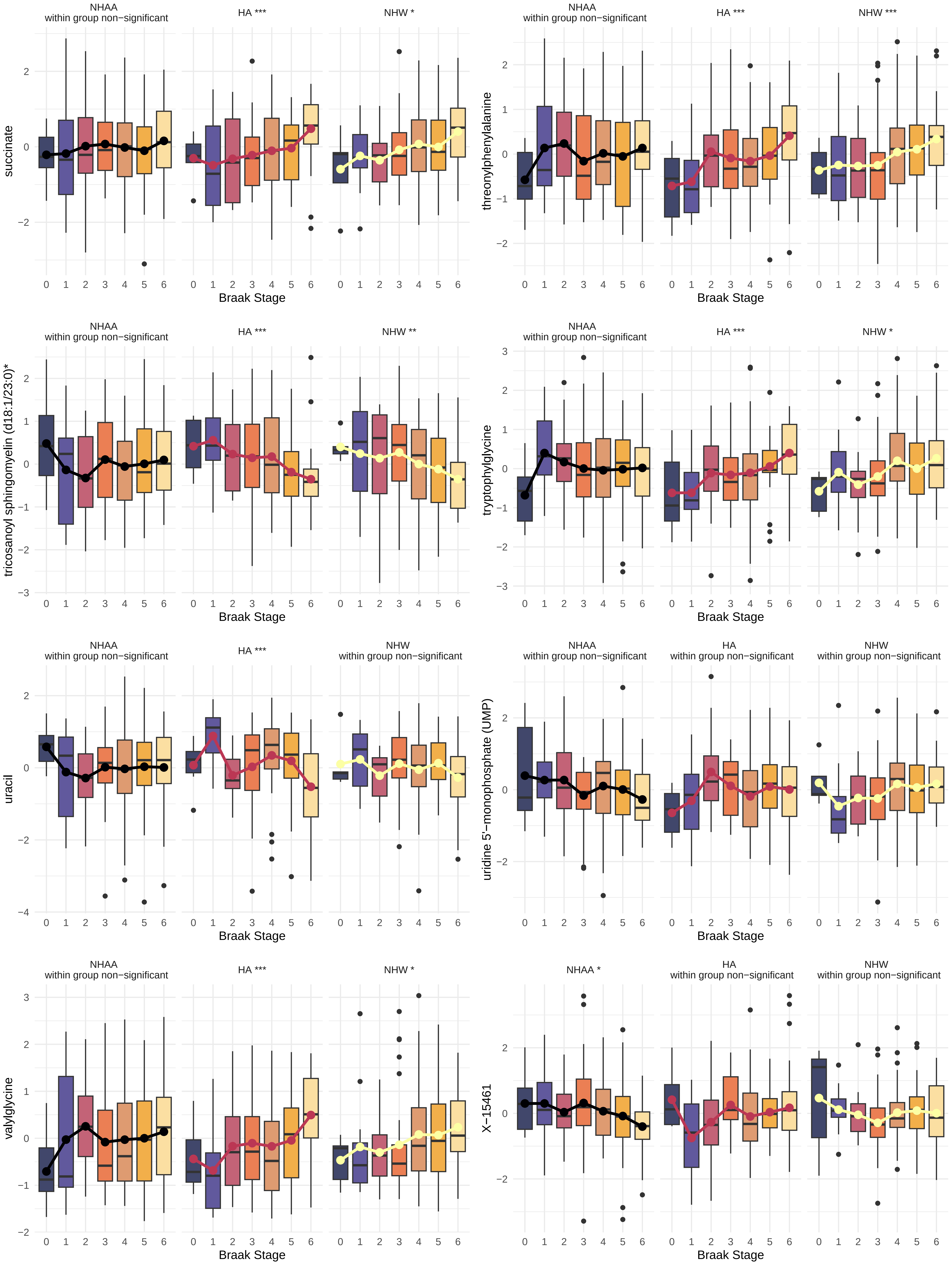

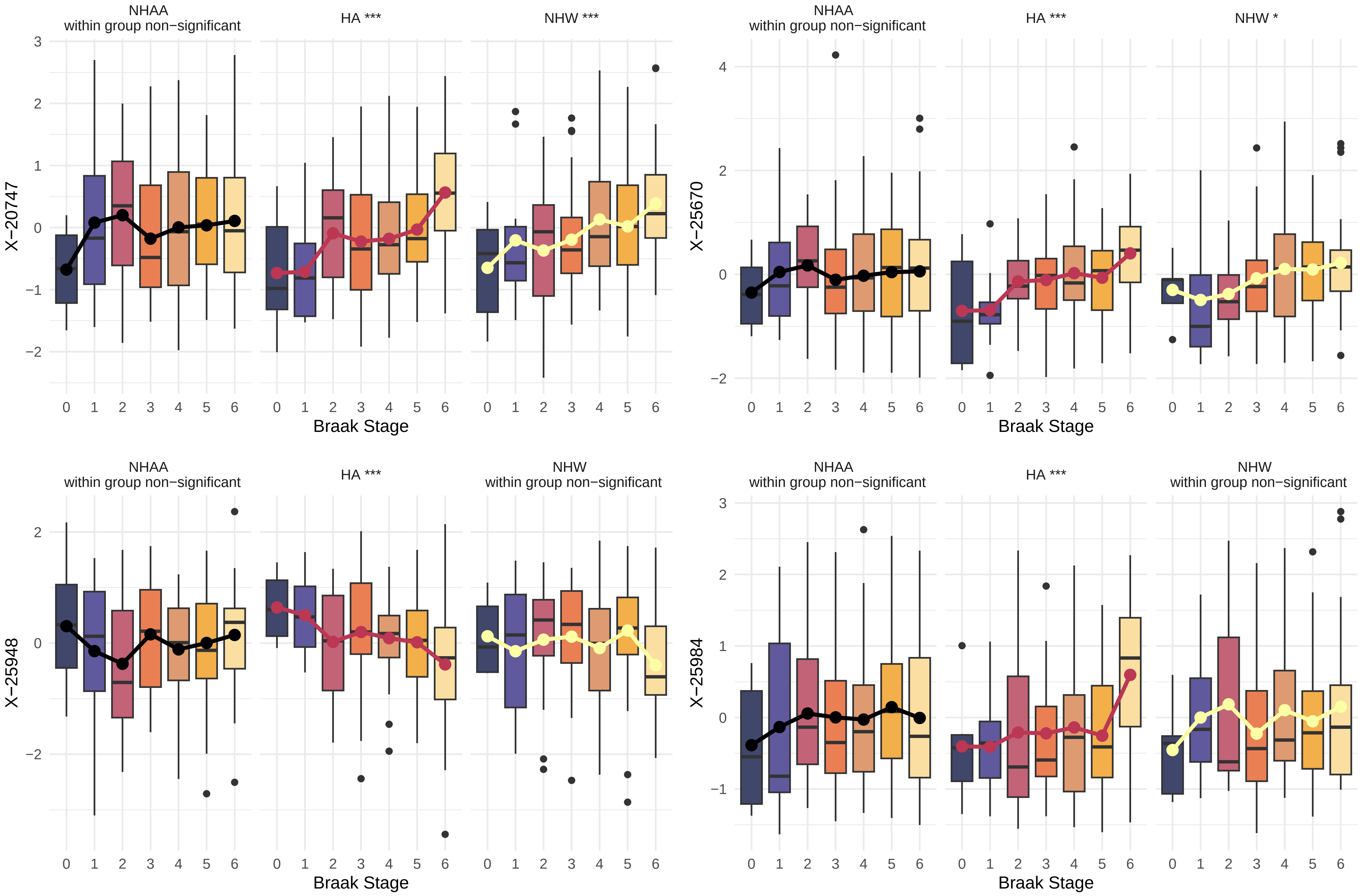
